# Applying Prospective Tree-Temporal Scan Statistics to Genomic Surveillance Data to Detect Emerging SARS-CoV-2 Variants and Salmonellosis Clusters in New York City

**DOI:** 10.1101/2024.08.28.24312512

**Authors:** Sharon K. Greene, Julia Latash, Eric R. Peterson, Alison Levin-Rector, Elizabeth Luoma, Jade C. Wang, Kevin Bernard, Aaron Olsen, Lan Li, HaeNa Waechter, Aria Mattias, Rebecca Rohrer, Martin Kulldorff

## Abstract

Genomic surveillance data are used to detect communicable disease clusters, typically by applying rule-based signaling criteria, which can be arbitrary. We applied the prospective tree-temporal scan statistic (TreeScan) to genomic data with a hierarchical nomenclature to search for recent case increases at any granularity, from large phylogenetic branches to small groups of indistinguishable isolates. Using COVID-19 and salmonellosis cases diagnosed among New York City (NYC) residents and reported to the NYC Health Department, we conducted weekly analyses to detect emerging SARS-CoV-2 variants based on Pango lineages and clusters of *Salmonella* isolates based on allele codes. The SARS-CoV-2 Omicron subvariant EG.5.1 first signaled as locally emerging on June 22, 2023, seven weeks before the World Health Organization designated it as a variant of interest. During one year of salmonellosis analyses, TreeScan detected fifteen credible clusters worth investigating for common exposures and two data quality issues for correction. A challenge was maintaining timely and specific lineage assignments, and a limitation was that genetic distances between tree nodes were not considered. By automatically sifting through genomic data and generating ranked shortlists of nodes with statistically unusual recent case increases, TreeScan assisted in detecting emerging communicable disease clusters and in prioritizing them for investigation.

## Introduction

Whole genome sequencing (WGS) data are increasingly used by public health officials for communicable disease surveillance and cluster detection (1). For example, SARS-CoV-2 variant surveillance allows officials to monitor the effects of new variants on COVID-19 disease severity, transmission, diagnostics, therapeutics, and immunity from prior infections and vaccinations (2, 3). In the U.S., variant data have guided decisions around COVID-19 vaccine composition and revocation of emergency use authorizations for monoclonal antibody therapies with decreased clinical efficacy (4). Such data have been used in New York City (NYC) to summarize epidemiologic characteristics of newly emerging variants (5–8), assess illness severity (9), and elucidate community transmission patterns (10). Timely knowledge of emerging variants with increased transmissibility or immune escape can prompt actions to limit spread. Such actions are particularly important in congregate settings and for populations at increased risk of severe illness, such as people who are older or living with comorbid conditions (11).

When many SARS-CoV-2 variants and recombinants cocirculate in a “swarm or soup of variants” (12), a key challenge is deciding in near-real time which ones to closely monitor over which time increments. Bioinformatic methods and phylodynamic models can be used to prioritize variants and estimate variant-specific growth rates (13, 14), although this can be onerous to operationalize with many cocirculating variants. In the COVID data tracker developed by the U.S. Centers for Disease Control and Prevention (CDC), lineages are displayed either if they account for >1% of sequences nationally during a 2-week period or have been classified as a variant of interest or concern (2, 15, 16).

WGS-based subtyping is also revolutionizing population-based enteric bacterial disease surveillance. When officials can quickly identify patients infected with genetically similar pathogens, the probability of identifying a common exposure and preventing further infections is increased (17, 18). The Public Health Laboratory (PHL) at the NYC Department of Health and Mental Hygiene (NYC Health Department) performs core genome multilocus sequence typing (cgMLST), using prescribed CDC PulseNet methods (19, 20). To detect *Salmonella* clusters, PHL staff compare cgMLST profiles of sequences stored in a local database of isolates (pathogens isolated from clinical specimens) tested and sequenced at PHL. This process often misses isolates sequenced out-of-jurisdiction, as data sharing can be difficult, and requires extensive manual input, which can lead to undetected clusters.

Enteric disease cluster detection is typically operationalized using static, rule-based definitions, in which isolates from patients in a geographic area are grouped within fixed cut- offs of genetic relatedness and time (21, 22). Rules can be established by using historical outbreak data and modeling (23, 24). A commonly used working cluster definition is ≥3 *Salmonella* isolates within a 60-day window within 10 alleles, where ≥2 cases are within ≤5 alleles (20, 23). Such rules, which can be arbitrary, vary across pathogens according to genetic diversity, ecology, and prevalence (25) and are more stringent for common serotypes, such as Enteritidis (26). Existing cluster detection tools (27–30) do not also analyze the extent that cases are spread out versus concentrated in time, despite the importance of temporal clustering for cluster detection and investigation.

In contrast, space-time scan statistics search flexibly in both space and time and can accommodate adjustments for purely temporal and purely spatial variation (31, 32). CDC and state and local health departments, including NYC, previously used a rule-based aberration detection method (the historical limits method (33–36)); CDC discontinued this approach in 2020 (37). In 2014, the NYC Health Department transitioned to using prospective scan statistics to quickly detect unusual clusters of any geographical size or duration for many reportable communicable diseases, including legionellosis, salmonellosis, and later COVID-19 (38–41).

Returning to WGS data, we wish to similarly search in a flexible manner in time. However, rather than searching flexibly in geographical location and size, we wish to be flexible in the location of patients’ WGS isolates on a phylogenetic tree and the granularity of nodes on that tree, to quickly detect increases at any single node or collection of closely related nodes.

Tree-temporal scan statistics (42, 43) are used by CDC, the U.S. Food and Drug Administration, and academic scientists to detect and evaluate unanticipated adverse reactions to pharmaceutical drugs and vaccines (44–50). In this pharmacovigilance context, potential adverse events can be classified in a tree structure based on *International Classification of Diseases, Tenth Revision* (ICD-10) diagnosis codes. The codes are grouped hierarchically, reflecting general or specific disease conditions affecting different body systems, with related diagnoses located on the same tree branch. Unusual increases in diagnoses at any level of specificity can be detected in sequential analyses, at any length of time after vaccine or drug administration.

Herein, we marry ideas of flexibly scanning prospectively in calendar time (as for spatiotemporal cluster detection) with flexibly scanning along a hierarchical tree structure (as is conducted for pharmacovigilance). We thereby establish an “innovation at the edge” of infectious disease epidemiology and pharmacoepidemiology (51). We introduce the prospective tree-temporal scan statistic and describe its real-time application by the NYC Health Department. We selected SARS-CoV-2 and *Salmonella* because of their substantial disease burdens (52) and availability of genomic surveillance data with a hierarchical nomenclature, with the potential to guide local public health actions.

## METHODS

### Genomic surveillance data

#### SARS-CoV-2

PHL and other laboratories perform WGS on a portion of specimens from confirmed COVID-19 cases (53) diagnosed among NYC residents, as previously described (5, 8, 54, 55). Weekly starting August 12, 2021, we determined counts of each lineage assignment during a rolling 12 week-period ending on the most recent specimen collection date. Pango lineages, which represent the dynamic nomenclature applied to genetically distinct SARS-CoV-2 lineages (56, 57), were assigned by using the pangolin software tool (58). Initially, we used the PangoLEARN machine learning model to assign a lineage name to each WGS result (58). To improve lineage assignment stability, we switched as of December 16, 2021 to the UShER method for placing new genome sequences onto a phylogeny (59, 60).

Occasionally, as with XBB.1.5 and then XBB.1.16, a variant newly emerged during the rolling 12-week study period and quickly became predominant. In these instances, so as not to obscure more recently emerging variants, we temporarily shortened the study period to begin after that variant became predominant, then returned to a rolling 12-week period. This is similar to an approach used to fine-tune a spatiotemporal cluster detection system when there is difficulty detecting new outbreaks in areas with recent prior outbreaks (41).

#### Salmonella

When a NYC resident tests positive for *Salmonella* infection, City and State laws require the laboratory to report the result and submit the patient’s isolate to PHL or the New York State Department of Health (61, 62). These laboratories conduct WGS on the isolates. WGS data (including serotype and cgMLST allele calls) are compiled with patient demographic data and uploaded to CDC PulseNet, where allele codes are assigned at the national level (20, 63). Allele codes are then populated in CDC’s System for Enteric Disease Response, Investigation, and Coordination (SEDRIC) (64). In parallel, graduate student interns at the NYC Health Department attempt to interview all NYC residents with salmonellosis as soon as feasible after initial report to collect possible exposure information (65).

Weekly from SEDRIC starting November 16, 2022, we downloaded allele codes for salmonellosis (typhoidal and nontyphoidal) for New York State residents, as additional parsing of patient addresses was necessary to restrict to NYC residents. Approximately 43% of the state’s residents live in NYC (66). We defined “NYC residents” as New York State residents known to live in NYC plus the <2% with unknown residency status, as we preferred to include a small number of non-NYC residents rather than potentially miss outbreaks involving NYC residents.

We determined counts of each *Salmonella* allele code during a rolling 365-day period ending on the most recent specimen collection date. Given delays between specimen collection and allele code assignment, we conducted weekly sensitivity analyses replacing the temporal element with the “upload date” in SEDRIC, with a rolling 365-day period ending with the most recent upload date.

#### Health equity

The population benefits of genomic surveillance might be inequitably distributed if particular groups are underrepresented in WGS results (67). Underrepresentation might be a consequence of inequitable access to health care and laboratory testing and, for SARS-CoV-2 infections, nonrandom sampling practices for sequencing (54). We assessed WGS result availability for confirmed and probable cases of COVID-19 (53) and salmonellosis (68) among NYC residents diagnosed during a 2-year period ending October 2023. We stratified by patient-level race or ethnicity and by the Index of Concentration at the Extremes, an area-based measure of economic and racial or ethnic segregation (69, 70).

### Hierarchical tree files

#### SARS-CoV-2

Pango lineage notes were used to determine parent-child relationships for all detected SARS-CoV-2 variants (58, 71). For example, for the analysis conducted on August 17, 2023, all detected variants were descended from B.1. Thus, B.1 was designated as the tree root, which progressively branched into increasingly specific lineages, including the Omicron variant (i.e., B.1.1.529), and culminating in more specific nodes, such as the Omicron subvariant EG.5.1.1. For recombinant lineages (e.g., XBB), we assigned multiple parents effective January 2024, but in the earlier analyses presented here, we assigned the most recent common ancestor as the parent (Table 1).

**Table 1.** Example hierarchical nomenclature for SARS-CoV-2 variants assigned to Pango lineages, showing tree levels.

| Level | Node (Pango lineage) | Note |
| --- | --- | --- |
| 1 | B.1 |  |
| 2 | B.1.1 |  |
| 3 | B.1.1.529 | Omicron |
| 4 | BA.2 | Alias of B.1.1.529.2 |
| 5 | XBB | Recombinant lineage of BJ.1 (alias of B.1.1.529.2.10.1.1) and BM.1.1.1 (alias of B.1.1.529.2.75.3.1.1.1) |
| 6 | XBB.1 |  |
| 7 | XBB.1.9 |  |
| 8 | XBB.1.9.2 |  |
| 9 | EG.5 | Alias of XBB.1.9.2.5 |
| 10 | EG.5.1 |  |
| 11 | EG.5.1.1 |  |

#### Salmonella

We designated “SAL” as the tree root and each *Salmonella* serotype (e.g., Typhi, Enteritidis, Kottbus) as the second tree level. We appended the allele code, which can be up to six digits, to the serotype. Whereas laboratory scientists typically compare isolates manually using allele ranges, we used allele codes because of the standardized hierarchical nomenclature. Isolates with more allele code digits in common have a lower number of allele differences (Table 2).

**Table 2.** Example hierarchical nomenclature for *Salmonella* isolates assigned a serotype and allele code.

| Level | Node (serotype, allele code) | Maximum expected allele difference for isolates matching at specified allele code digit |
| --- | --- | --- |
| 1 | SAL | N/A |
| 2 | SAL.Kottbus | N/A |
| 3 | SAL.Kottbus.6185 | 80 |
| 4 | SAL.Kottbus.6185.1 | 28 |
| 5 | SAL.Kottbus.6185.1.1 | 15 |
| 6 | SAL.Kottbus.6185.1.1.2 | 7 |
| 7 | SAL.Kottbus.6185.1.1.2.1 | 4 |
| 8 | SAL.Kottbus.6185.1.1.2.1.1 | 0<sup>a</sup> |

N/A = not applicable.
<sup>a</sup>Isolates matching at the sixth digit of the allele code are 0 alleles apart, i.e., indistinguishable by core genome multilocus sequence typing.

### Prospective tree-temporal scan statistic

To flexibly search for unusual increases in any variant or allele code emerging over any recent time period, we conducted weekly prospective analyses using the tree-temporal scan statistic (42, 72) using the free TreeScan^TM^ software (73). We conditioned analyses on time, representing the specimen collection date, to adjust nonparametrically for any citywide purely temporal patterns, such as data reporting lags or increasing or decreasing trends. We also conditioned on “node,” representing the variant or allele code, to account for whether cases historically had been common or rare at each node during the baseline period (i.e., prior to the cluster period). This is because we were interested in detecting newly emerging nodes, not nodes that were also common during the baseline period. Mathematical formulae for calculating the expected number of cases, excess cases, and relative risk are available in the TreeScan user guide (72).

We set the prospective tree-temporal scan statistic to detect emerging disease clusters at any node in the WGS-based tree file beyond the root, from large phylogenetic branches to small groups of genetically indistinguishable isolates. For SARS-CoV-2, we searched for increases in variants during the most recent 14, 15, 16, …, 27, or 28 days to balance recency and persistence. For *Salmonella*, we searched for allele codes with increases during the most recent 1, 2, 3, …, 89, or 90 days to encompass the standard 60 days in the rule-based *Salmonella* definition (20), plus an additional 30 days to accommodate data lags.

For each node (candidate cluster), a likelihood ratio-based test statistic is calculated when the observed number of cases during the time window at the node exceeds the expected number. The candidate cluster with the maximum likelihood ratio test statistic is the cluster least likely to be due to chance under the null hypothesis of no node-by-time interaction, after adjusting for purely temporal variation and total node counts during the study period. For example, if a node has 5.4% of cases during the baseline period, and there are 100 total cases with WGS results during the cluster period, then the expected number of cases during the cluster period at that node is 5.4.

Monte Carlo hypothesis testing is used to assess statistical significance, controlling for the multiplicity of overlapping nodes and time windows evaluated. To create a simulated dataset, case dates are shuffled and randomly assigned to the original nodes. The maximum likelihood ratio test statistic for each simulated dataset is calculated in the same way as for the observed dataset. The standard number of Monte Carlo replications is 999, but to slightly improve performance, we used 99,999 for *Salmonella* analyses and 999,999 for SARS-CoV-2 analyses, which had fewer tree nodes.

The maximum likelihood ratio for the observed dataset is ranked among the ones from the simulated runs under the null hypothesis, and a *P* value is derived from this ranking as *P*=rank/(999,999+1) for the SARS-CoV-2 analyses (72). For prospective analyses, a recurrence interval (RI) is calculated as the reciprocal of the *P* value. For a weekly analysis frequency, this is further divided by 52 for the number of analyses per year. The RI represents the duration of weekly surveillance required for the expected number of clusters at least as unusual as the observed cluster to be equal to 1 by chance (74). For example, when the null hypothesis of no clusters is true, then during a 1-year period, the expected number of clusters with RI ≥ 365 is 1.

We defined a signal as any cluster with RI ≥100 days for *Salmonella* or ≥365 days for SARS-CoV-2. We considered RI 100–<365 days as a weak cluster, RI 365 days–<5 years as a moderate cluster, RI 5–<100 years as a strong cluster, and RI ≥100 years as a very strong cluster (41). Web Appendix 1 provides weekly cluster reporting details.

### Performance assessment

#### SARS-CoV-2

In the absence of national guidance for how jurisdictions should select variants to monitor locally, we compiled illustrative examples of successes and challenges in using TreeScan results to focus attention on emerging variants during weekly analyses conducted during August 2021–November 2023.

#### Salmonella

We characterized clusters prospectively detected by TreeScan during the first year of weekly analyses, November 16, 2022–November 8, 2023. We considered clusters to be “solved” if investigators identified a common food source, animal exposure, exposure site, or travel history that likely explained the association among cluster patients. Web Appendix 2 provides further details about cluster definitions, cluster prioritization, and consideration of typhoidal clusters.

### Ethics statement

The Institutional Review Board of the NYC Health Department determined this activity meets the definition of public health surveillance as set forth under 45 CFR§46.102(l)(2).

## RESULTS

### Completeness and representativeness

Among NYC residents diagnosed during November 2021–October 2023, WGS was conducted for 7% of COVID-19 cases (151,944 of 2,266,600) and for 62% of nontyphoidal salmonellosis cases (1,679 of 2,722; Web Table 1). Of 1,068 salmonellosis cases with no allele code, 937 (88%) were probable cases and positive only by culture-independent diagnostic testing, 106 (10%) were culture-positive but had no isolate available for WGS, 5 (<1%) underwent WGS but failed quality control, and 20 (2%) were unique sequences that could not be matched to an existing allele code by CDC’s naming algorithm. The median lag from specimen collection to allele code assignment was 22 days (interquartile range: 20–28 days). Of 1,654 salmonellosis cases with an allele code assigned, 1,322 (80%) had a fully or partially completed interview; interviews are necessary to collect information for identifying common exposures among cluster patients.

Patient demographic characteristics were similarly distributed between reported cases overall and the subset with WGS results. Distributions were within +/-2.5% for every stratum of race or ethnicity and the Index of Concentration at the Extremes (Web Table 1). Although substantial portions of patients lacked WGS results, there was no evidence of systematic underrepresentation during this period.

### SARS-CoV-2 illustrative examples

#### Rapid detection of a locally emerging variant

The weekly analysis performed June 22, 2023, with a computer running time of 4 minutes and 15 seconds, identified 6 SARS-CoV-2 variants emerging among NYC residents (Table 3). Three of the 6 nodes were the same as in the prior week’s analysis (Web Table 2), including persistent, strong signals for XBB.1.16, XBB.2.3, and their subvariants. Of the newly signaling nodes, EG.5.1 (RI = 35 years) first signaled more strongly than its grandparent (XBB.1.9.2), with 11 specimens collected during May 17–June 12, 2023. EG.5 or EG.5.1 continued to signal for 13 consecutive weekly analyses, June 22– September 14, 2023, after which more specific subvariants (e.g., EG.5.1.6) began signaling more strongly.

**Table 3.** Analysis conducted on June 22, 2023 to apply prospective tree-temporal scan statistics to detect emerging SARS-CoV-2 variants in specimens collected among New York City residents during the 14–28-day period ending June 12, 2023.

| Variant | No. of cases with specimens collected during 12-week study period, March 21–June 12, 2023 | Cluster start date, ending June 12, 2023 | No. of cases with specimens collected during cluster window | No. of expected cases | Relative risk | No. of excess cases | Test statistic | Recurrence interval (years)<sup>b</sup> | No. of consecutive weeks signaling | Percent of sequenced cases with specimens collected week ending June 12, 2023 |
| --- | --- | --- | --- | --- | --- | --- | --- | --- | --- | --- |
| XBB.1.16<sup>a</sup> | 213 | May 16 | 107 | 52.1 | 3.7 | 78.0 | 22.8 | 19,231 | 11 | 28 |
| XBB.2.3<sup>a</sup> | 83 | May 18 | 41 | 17.5 | 3.9 | 30.5 | 11.5 | 4,808 | 3 | 17 |
| XBB.1.24.1 | 5 | May 30 | 5 | 0.4 | ∞ | 5.0 | 8.2 | 55 | 1 | <1 |
| EG.5.1<sup>c</sup> | 11 | May 17 | 11 | 2.5 | ∞ | 11.0 | 7.9 | 35 | 1 | 6 |
| XBB.1.5.68 | 8 | May 19 | 8 | 1.6 | ∞ | 8.0 | 6.6 | 5 | 1 | <1 |
| XBB.1.5.16<sup>a</sup> | 13 | May 16 | 11 | 3.2 | 17.3 | 10.4 | 5.8 | 2 | 2 | <1 |

<sup>a</sup>Subvariants included.
<sup>b</sup>Nodes with recurrence interval $\geq 1$ year were included. The maximum possible recurrence interval for this analysis was 19,231 years. When using 999,999 Monte Carlo replications, the smallest possible $P$ -value is $1/999999 = 0.000001$ . With a weekly prospective analysis frequency, the maximum recurrence interval was thus $(1 / 0.000001) / 52$ analyses per year = 19,231 years.
<sup>c</sup>Alias of XBB.1.9.2.5.1.

WHO designated EG.5 as a variant under monitoring on July 19, 2023 and a variant of interest on August 9, 2023 (75), 4 and 7 weeks, respectively, after our first EG.5.1 signal. During a period with many cocirculating variants and when EG.5 initially constituted a small number and percentage of cases with WGS results, the TreeScan analysis and trend visualization (Figure 1) led NYC Health Department officials to focus attention on this variant.

**Figure 1.**
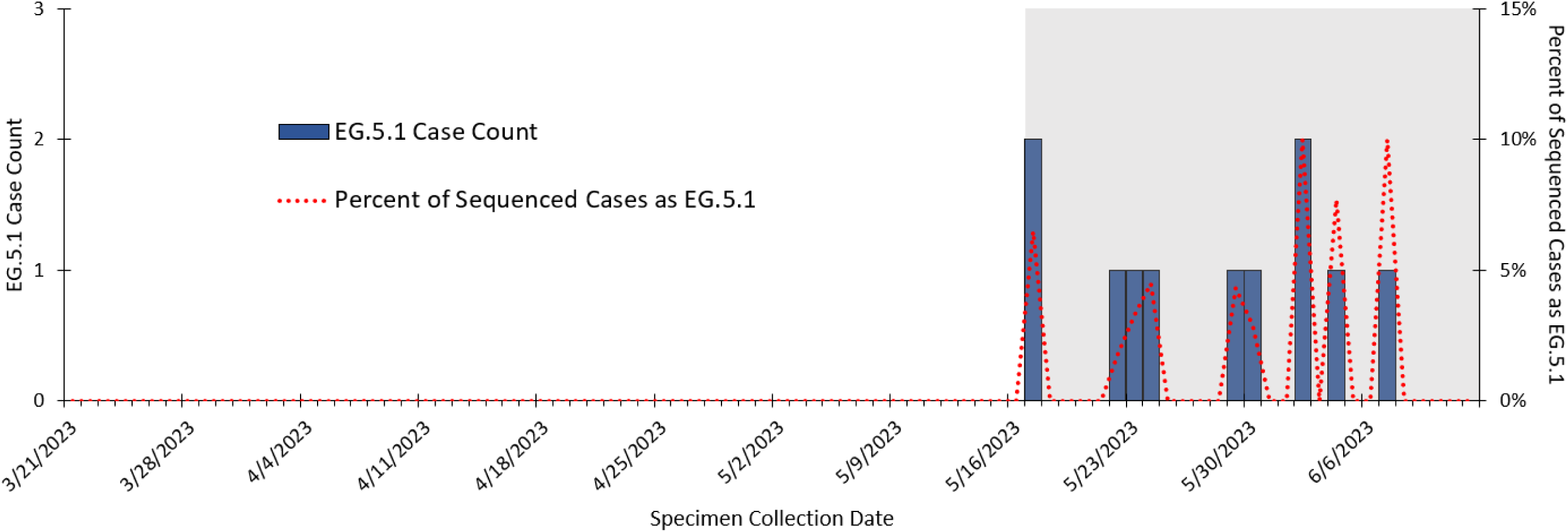
Count and percent of cases with EG.5.1 as the SARS-CoV-2 sequencing result among New York City residents with specimens collected during March 21–June 12, 2023. Results are as of June 22, 2023, the first analysis week that EG.5.1 signaled as emerging, with the cluster window starting May 17, 2023 shaded in grey. *Alt text for accessibility*: A time series graph showing 0 cases of the SARS-CoV-2 variant EG.5.1 until May 17, 2023, followed by 11 cases during May 17 through June 7, 2023, with 0, 1, or 2 cases per day.

#### Delayed detection of locally emerging variants

In the analysis performed October 20, 2022, multiple BE.1.1.1 subvariants first signaled as having emerged since September 19, 2022, indicating delayed detection of BQ.1, BQ.1.1 (RI = 19,231 years for both nodes), and BQ.1.3 (RI = 2.4 years). In the concurrent UShER version update, a subset of cases had been reassigned to BQ lineages, revealing that BQ lineages, which descended from BE.1.1.1 (71), had been present for >1 month. Once the input data were updated, TreeScan analyses appropriately detected the emergence of BQ lineages.

#### Assurance of no other locally emerging variants

The Omicron variant was first detected in NYC in clinical and wastewater samples collected in November 2021 and quickly became predominant (54, 76). While staff were urgently focused on characterizing local effects on population health, TreeScan analyses provided assurance there were no additional lineages emerging concurrently that also would have required attention and response.

#### Salmonella

During the first year of weekly analyses, on 128 serotypes, TreeScan detected 16 unique clusters in the primary analysis using specimen collection date as the temporal element, and 1 additional cluster in the sensitivity analysis using upload date (Table 4). TreeScan detects statistical anomalies, which must be investigated to distinguish true clusters from data quality issues. Of the 17 clusters, 2 represented data quality issues that were quickly resolved in SEDRIC, where >1 isolate had been sequenced from the same patient. The remaining 15 clusters were credible and worth investigating. Of these 15 clusters, 2 were typhoidal and associated with travel to an endemic area. Of the 13 nontyphoidal clusters, 2 (15%) were comprised of family members with shared exposures, 2 (15%) reflected larger, interjurisdictional outbreaks, 2 (15%) were persistent strains causing illnesses over a long time (77–80), and 7 (54%) were unsolved. Of the 13 nontyphoidal clusters, 1 (8%) was detected at the third allele code digit so encompassed a broader allele range than rule-based cluster definitions. The remaining 12 (92%) were detected to at least the fifth allele code digit, aligning with rule-based cluster definitions. Of these clusters, 9 (75%) were concurrently detected by PHL, 2 (17%) were comprised entirely of isolates tested at other jurisdictions’ public health laboratories so could not have been detected by PHL, and 1 (8%) was detected when technological issues disrupted portions of PHL’s cluster detection workflow.

**Table 4.** Salmonellosis clusters among New York City residents detected using prospective tree-temporal scan statistics in weekly analyses conducted during November 16, 2022–November 8, 2023.

| Serotype | Allele code | Date first detected | Cluster start date (specimen collection date) | Observed cases | Relative risk | Excess cases | Recurrence interval | Notes |
| --- | --- | --- | --- | --- | --- | --- | --- | --- |
| Typhimurium | 6745.56.1.2.3.116 | 12/8/22 | 10/25/22 | 4 | ∞ | 4.0 | 7.5 years | Unsolved |
| Oranienburg | 6760.67.167.5.1.1 | 12/21/22 | 11/19/22 | 2 | ∞ | 2.0 | 130 days | Data quality issue: removed duplicate isolate |
| Kottbus | 6185.1.1.2.1 | 1/4/23 | 12/1/22 | 5 | 25.6 | 4.8 | 1.9 years | Unsolved |
| I 4:i:- | 772.1.3.1.2.2 | 1/11/23 | 12/8/22 | 3 | ∞ | 3.0 | 2.9 years | Unsolved |
| Typhimurium | 6745.20.1.1.10.9 | 2/1/23 | 1/9/23 | 3 | ∞ | 3.0 | 12 years | Same family. Multiple shared exposures |
| Typhimurium | 6766.47.1.12.4.3 | 3/1/23 | 2/10/23 | 2 | ∞ | 2.0 | 33 years | Unsolved |
| Typhi | 6788.1.1.47.1.97 | 3/15/23 | 2/15/23 | 4 | ∞ | 4.0 | 481 years | Same family. All traveled to endemic area |
| Paratyphi A | 1082.1.3.1.1.11 | 4/19/23 | 3/24/23 | 2 | ∞ | 2.0 | 119 days | Same family. All traveled to endemic area |
| Typhi | 6788.1.1.4.18 | 5/3/23 | 4/10/23 | 2 | ∞ | 2.0 | 147 days | Data quality issue: removed isolate from a follow-up specimen |
| Typhimurium | 6766.32.1.3.8.3 | 5/17/23 | 4/28/23 | 2 | ∞ | 2.0 | 339 days | Unsolved |
| Typhimurium | 459.3.1.8.1.1 | 6/7/23 | 5/15/23 | 2 | ∞ | 2.0 | 174 days | Same family. Shared meal |
| Paratyphi B<br>Var. L(+)<br>Tartrate+ | 316.6.3.13.1 | 7/19/23 | 7/4/23 | 2 | ∞ | 2.0 | 31 years | Unsolved |
| Javiana | 6765.1.1 | 9/7/23 | 7/21/23 | 7 | 14.6 | 6.5 | 213 days | 3 subgroups of more closely related isolates, 2 of which were multistate investigations |
| Infantis | 6747.16.3.109.1 | 9/13/23 | 7/28/23 | 4 | ∞ | 4.0 | 101 days | Multistate investigation |
| IIIb 61:k:1,5 | 135.5.33.6.1.1 | 9/27/23 | 9/26/23<sup>a</sup> | 2 | ∞ | 2.0 | 114 days | Unsolved |
| Newport | 6809.9.1.1.1.954 | 10/12/23 | 9/24/23 | 2 | ∞ | 2.0 | 6.2 years | Persisting enteric bacterial strain |
| Hadar | 6771.1.1.30.1 | 11/8/23 | 10/17/23 | 3 | 126.1 | 3.0 | 2.6 years | Persisting enteric bacterial strain |

<sup>a</sup>This was the only unique cluster detected in sensitivity analyses, where the temporal element was upload date instead of specimen collection date.

Investigators considered the TreeScan results to be helpful in focusing staff attention and investigation resources. For example, TreeScan clusters in NYC occasionally reflected concurrent, aberrant clusters across other jurisdictions, prompting multijurisdictional collaboration to identify common exposures. The TreeScan cluster at the third allele code digit alerted investigators to a local increase in *Salmonella* Javiana, spurring further investigation into possible subclusters. Moreover, in analyzing isolates according to location of residence, TreeScan detected clusters among NYC residents that otherwise might not have been detected because patients were tested by different laboratories. TreeScan also provided coverage when external technological issues disrupted certain processes at PHL.

## DISCUSSION

We introduced the prospective tree-temporal scan statistic, which, when applied to genomic surveillance data with a standardized hierarchical nomenclature, automatically sifts through large quantities of data in minutes and generates a ranked shortlist of nodes with a statistically unusual number of recent cases. Our method flexibly evaluates all candidate clusters, across many degrees of genetic relatedness and date ranges. It dynamically accounts for any purely temporal trends, such data lags or changes in WGS result availability, and minimizes false signals by adjusting for the multiplicity of nodes and cluster windows scanned. With real-time application to SARS-CoV-2 and *Salmonella* data, the NYC Health Department detected credible clusters for investigation and data quality problems for correction.

## Limitations

WGS results were available for only 7% of COVID-19 cases and 62% of salmonellosis cases. After the federal COVID-19 public health emergency declaration ended in May 2023 and with reduced funding, specimen and sequence availability have declined (81), which could reduce population representativeness and delay new variant detection. Additionally, although the NYC Health Code requires laboratories to reflexively culture certain enteric pathogens, including *Salmonella* (61), the widespread use of culture-independent diagnostic testing has reduced the proportion of salmonellosis cases with recovered isolates. Patients without WGS results cannot contribute their exposure histories to WGS cluster investigations, making it more challenging to solve outbreaks. Improving population-based WGS data completeness, representativeness, and timeliness requires strengthening partnerships with clinics and submitting laboratories and deepening investments in laboratory capacity and bioinformatics infrastructure, including applying culture-independent sequencing methods (82–85).

Where WGS results were available, the tree nomenclature imposed limitations. For SARS-CoV-2, assigning Pango lineages using UShER allowed for accurate and stable lineage assignments at the expense of timeliness. As in the BQ lineages example, delays in updating nomenclature to recognize new lineage designations resulted in delayed detection. For *Salmonella*, the signal-to-noise ratio to detect a new cluster is poor for allele codes ending in “x,” such as “SALM1.0 – 6743.2.4x” for the serotype Enteritidis (26), as the underlying 4-, 5-, and 6-digit codes are masked due to low genomic diversity increasing the within-code distance beyond assignment thresholds. More broadly, we rely on a standardized nomenclature, with no consideration of genetic distances between tree nodes.

TreeScan should complement, not replace, other cluster detection approaches using laboratory-based data, such as by examining allele ranges (20, 23). Tree-temporal scan statistics could miss outbreaks where genetic or temporal clustering is weak. Zoonotic disease outbreaks, such as those associated with exposure to reptiles or backyard poultry, often involve multiple serotypes with large allelic diversity (25, 86). Patients’ isolates might be weakly clustered temporally for outbreaks due to persistent environmental contamination (87) or following delays in accessing medical care or obtaining WGS results. Web Appendix 3 provides additional minor limitations.

## Conclusions

By decreasing reliance on time-consuming, manual laboratory data review, and by simultaneously analyzing data not only by genetic relatedness but also by temporal clustering, TreeScan analyses can help officials focus limited investigative resources on emerging clusters and variants. Future work could apply this approach to additional pathogens (20, 88, 89) using additional hierarchical nomenclature systems (e.g., SNP addresses (90)), analyze additional pathogen characteristics (e.g., antimicrobial resistance patterns), and analyze state- and national-level data to support multijurisdictional outbreak response. Incorporating TreeScan into analytical pipelines could strengthen strategic frameworks for genomic surveillance (91), including in low- and middle-income countries (92).

Health departments should continuously apply multiple cluster detection methods to quickly detect different types of outbreaks. In NYC, building-level analyses and spatiotemporal scan statistics have quickly detected outbreaks with strong geographic clustering, before laboratory subtyping results became available (39, 93). However, these methods could miss geographically diffuse outbreaks, such as following exposure to a widely disseminated source, or outbreaks affecting only a few patients. Despite lags in subtyping data availability, such outbreaks could be detected faster by applying tree-temporal scan statistics to WGS data.

TreeScan thus fills an important gap in the public health practitioner’s automated cluster detection and monitoring toolkit.

## Supporting information

Web Material

## Data Availability

Data, software, and code are available as follows at the links below: (1) SARS-CoV-2 variant data for New York City residents. (2) Allele codes for Salmonella isolates (available to CDC partners via SEDRIC). (3) SAS code for generating TreeScan input files. (4) TreeScan software for free download. (5) TreeScan source code.

https://github.com/nychealth/coronavirus-data/tree/master/variants

https://www.cdc.gov/foodborne-outbreaks/php/foodsafety/tools/

https://github.com/CityOfNewYork/communicable-disease-surveillance-nycdohmh

https://www.treescan.org/

https://github.com/scanstatistics/treescan

## Acknowledgments

This work was supported by the U.S. Centers for Disease Control and Prevention (NU90TP922035-05, NU50CK000517-01-09, NU50CK000517-05-00).

SARS-CoV-2 variant data for NYC residents are available on GitHub (<u>github.com/nychealth/coronavirus-data/tree/master/variants</u>). Allele codes for *Salmonella* isolates are available to CDC partners via SEDRIC (https://www.cdc.gov/foodborne-outbreaks/php/foodsafety/tools/). SAS code for generating TreeScan input files is available on GitHub (<u>github.com/CityOfNewYork/communicable-disease-surveillance-nycdohmh</u>). The TreeScan software (www.treescan.org) and source code (<u>github.com/scanstatistics/treescan</u>) are freely available.

The authors thank Scott Hostovich at Information Management Services, Inc. for incorporating updates to the TreeScan software. We thank Helly Amin, Ahmed Rahat, Dr. Faten Taki, Olivia Samson, Naama Kipperman, and Dr. Mustapha Mustapha for analytic contributions. Mark Alexander and Kuan Chen set up SQL databases for automated data transfers. We thank Team *Salmonella* for exceptional work to complete timely patient interviews, which is key to identifying outbreak sources, as well as Lenka Malec, Marisa Gerard, Danielle Martinez, Samuel Davey, John Croft, and Athanasia Papadopoulos for managing and leading numerous salmonellosis cluster investigations. Christian King, Wai Sum So, Bun Tha, Moinuddin Chowdhury, and Nelson De La Cruz worked tirelessly in PHL’s Molecular Typing Laboratory. We thank Vasudha Reddy for sustained support and guidance on these activities. We thank Wadsworth Center for sequencing *Salmonella* isolates from NYC residents that were not sent to PHL. We thank CDC PulseNet database managers for assigning allele codes and Dr. Lyndsay Bottichio and the Surveillance, Information Management, and Statistics Office in CDC’s Division of Foodborne, Waterborne, and Environmental Diseases for SEDRIC data provision. We thank CDC FoodCORE for continued funding to support both epidemiology and laboratory capacity for foodborne disease surveillance in NYC. Aviva Goldstein, Lauren da Fonte, and the Fund for Public Health in New York City provided fundraising and grant management support for TreeScan. Drs. Judy Maro and Katherine Yih provided valuable suggestions on an early manuscript draft.

A preliminary version of this work was presented at the Integrated Foodborne Outbreak Response and Management (InFORM) Virtual Conference, April 26–29, 2022.

## Abbreviations

CDC, Centers for Disease Control and Prevention; cgMLST, core genome multilocus sequence typing; ICD-10, *International Classification of Diseases, Tenth Revision*; NYC, New York City; PHL, Public Health Laboratory, NYC Department of Health and Mental Hygiene; RI, recurrence interval; SEDRIC, System for Enteric Disease Response, Investigation, and Coordination; WGS, whole genome sequencing.

