## Supplementary material for "Applying Prospective Tree-Temporal Scan Statistics to Genomic Surveillance Data to Detect Emerging SARS-CoV-2 Variants and Salmonellosis Clusters in New York City": Web Material

Table of Contents


| | | |
|---|---|---|
| Web Appendix 1 | Weekly reporting of TreeScan cluster results. | P. 2 |
| Web Appendix 2 | Details about cluster definitions for genomic surveillance of *Salmonella* data applied by the NYC Health Department, cluster prioritization, and consideration of typhoidal clusters. | P. 3 |
| Web Appendix 3 | Minor limitations of applying prospective tree-temporal scan statistics for cluster detection. | P. 4 |
| Web Table 1 | Whole genome sequencing result availability for confirmed and probable COVID-19 and nontyphoidal salmonellosis cases diagnosed during November 2021–October 2023 among New York City residents. | P. 6 |
| Web Table 2 | Analysis conducted on June 15, 2023 to apply prospective tree-temporal scan statistics to detect emerging SARS-CoV-2 variants in specimens collected among New York City residents during the 14–28-day period ending May 31, 2023. | P. 7 |

**Web Appendix 1.** Weekly reporting of TreeScan cluster results.

Analysts prepared internal weekly cluster reports. For overlapping signaling nodes, we typically selected the node with the highest likelihood ratio. At times, we instead selected a child node for greater genomic specificity despite a lower likelihood ratio, if the child node accounted for almost all cases during the cluster window for the parent node. For *Salmonella* at times, we instead selected a parent node with a few additional cases despite a lower likelihood ratio, since reviewing the additional patients' exposures could be informative.

SARS-CoV-2. For each signaling variant, the report indicated the number of consecutive weeks signaling, the percentage of sequenced cases among NYC residents for that variant as of the most recent week, the parent node, any child nodes also signaling, and notes regarding distinguishing mutations, prevalence, growth advantage, immune escape, and disease severity, if known. We also provided temporal graphs for each variant and a tree visualization, highlighting signaling nodes. National and regional estimated variant percentages from CDC's COVID data tracker (1) were included for context.

Signaling nodes were included in a monthly report for NYC Health Department leadership. This report supported situational awareness for emerging variants regarding temporal trends, geographic distribution, and proportions of patients, by variant, who were hospitalized or died.

*Salmonella*. For each signaling allele code, we notified investigators and generated a case line list. For all clusters detected by TreeScan or other methods, investigators attempted to identify possible common exposures, including by prioritizing interview attempts, reviewing patient demographic and interview data, and searching in SEDRIC to coordinate with other jurisdictions with related cases. If a common exposure source was identified, additional investigation activities could include environmental sampling or requesting shopper card data or food purchase receipts.

**Web Appendix 2.** Details about cluster definitions for genomic surveillance of *Salmonella* data applied by the NYC Health Department, cluster prioritization, and consideration of typhoidal clusters.

Although CDC PulseNet recommends defining a cluster as ≥3 clinical isolates (2, 3), the Public Health Laboratory (PHL) at the NYC Health Department applies a lower threshold of ≥2 isolates to avoid missing outbreaks. This is consistent with a prior CDC PulseNet cluster definition, before WGS became predominant, of ≥2 isolates within 60 days with indistinguishable pulsed-field gel electrophoresis patterns, if above baseline (3). PHL applies a rule-based definition of ≥2 isolates within a 60-day window within 0 alleles for *Salmonella* Enteritidis, within 5 alleles for *Salmonella* Typhimurium, and within 10 alleles for all other serotypes.

NYC Health Department epidemiologists review data for patients in all salmonellosis clusters meeting these rule-based criteria. Although all clusters are investigated, criteria that might increase epidemiologists' concern for further scrutiny include clusters with a rare serotype, a higher case count, a shorter time span, a smaller geographic span, inclusion in a multistate investigation, or matches with non-clinical isolates. Prior to implementing TreeScan analyses, statistical criteria were not considered in WGS cluster prioritization.

Of note, NYC has among the highest incidence of typhoid and paratyphoid fever in the U.S. (4). Nearly all such patients traveled to endemic areas, typically visiting friends and relatives in South Asia (5). Travel history is ascertained via patient interviews or health care provider reporting, and solving clusters by identifying a common country or region of travel does not affect local public health practice. To strengthen generalizability to jurisdictions where a larger proportion of salmonellosis is nontyphoidal, we distinguished typhoidal and nontyphoidal clusters in the results section of the main text.

**Web Appendix 3.** Minor limitations of applying prospective tree-temporal scan statistics for cluster detection.

First, not all aspects of cluster detection are fully automatable. A limitation of scanning across a hierarchical nomenclature is that clusters are nested as opposed to discrete (6). Epidemiologic judgment is required to select which of multiple, closely overlapping nodes to report and investigate. For SARS-CoV-2, whether one or more lineages are emerging more strongly than the parent lineage can be ambiguous. For *Salmonella*, choosing whether to investigate a signaling allele code that is more specific with fewer cases versus less specific with more cases might depend not only on cluster statistics, but also on investigation capacity at the time. If certain tree levels are not of interest, such as increases at serotype resolution (i.e., tree level 2 in Table 2), then users can apply the “Do not evaluate tree levels” TreeScan option, which could reduce the number of signals for manual inspection.

Second, TreeScan is useful for *early* cluster detection, but not for *prediction* of which clusters will continue to demonstrate excess growth (7). At first signal detection, nodes for which, in retrospect, case increases will prove to be temporary might be statistically indistinguishable from nodes that will proceed to grow into major outbreaks. Information external to TreeScan can be used for prediction, such as by assessing the genetic mutation profile and growth advantage of signaling nodes.

Third, historic outbreaks can interfere with subsequent outbreak detection. Cases in an historic outbreak will increase the recent expected case count such that it is harder to detect a new outbreak at that node and higher tree levels. This is only an issue for the duration of the study period. For example, with a rolling 365-day study period for salmonellosis, outbreaks that occurred >365 days prior cannot influence the baseline. One could also exclude days during major previous outbreaks from the baseline period in the input file (8).

Fourth, by conditioning analyses on both node and time, we adjusted for citywide data lags. If data lags were not a major issue, then an alternative analysis would be to condition on node and apply a time-uniform probability model.

Finally, formal, quantitative evaluation of new cluster detection methods can be challenging in public health practice. Salmonellosis clusters might not be “solved” for many reasons (9), unrelated to the quality of cluster detection methods. These reasons include whether patients agree to be interviewed, whether interviewed patients recall an accurate acute illness onset date, whether interviewed patients are reliable historians of their exposures, whether the outbreak source is common or rare, and whether the outbreak affects many patients or is multijurisdictional such that there are additional lines of evidence to triangulate exposures. Unlike in artificial data simulation environments (10), in practice, there is no gold standard of true outbreaks, only those that are detected and subjectively considered to be worth investigating. Future TreeScan evaluations could consider process-based metrics, such as person-hours saved in detecting credible clusters, together with outcomes-based metrics, such as the percentage of clusters solved.

**Web Table 1**. Whole genome sequencing result availability for confirmed and probable COVID-19 and nontyphoidal salmonellosis cases diagnosed during November 2021–October 2023 among New York City residents.

| Level | Stratum | COVID-19 | | Nontyphoidal salmonellosis | |
|---|---|---|---|---|---|
| | | No. cases (%) | No. with SARS-CoV-2 variant result (%) | No. cases (%) | No. with isolates available for sequencing (%) |
| | Total | 2,266,600 | 151,944 | 2,722 | 1,679[a] |
| Race or ethnicity | Asian/Pacific Islander | 292,287 (12.9%) | 21,491 (14.1%) | 441 (16.2%) | 276 (16.4%) |
| | Black/African American | 355,045 (15.7%) | 26,375 (17.4%) | 329 (12.1%) | 221 (13.2%) |
| | Hispanic/Latino | 588,882 (26.0%) | 38,237 (25.2%) | 798 (29.3%) | 500 (29.8%) |
| | White | 588,944 (26.0%) | 39,146 (25.8%) | 620 (22.8%) | 365 (21.7%) |
| | Another group | 18,816 (0.8%) | 1,455 (1.0%) | 127 (4.7%) | 81 (4.8%) |
| | Unknown | 422,626 (18.6%) | 25,240 (16.6%) | 407 (15.0%) | 236 (14.1%) |
| Index of Concentration at the Extremes[b] | Quintile 1 (least privileged) | 422,737 (18.7%) | 28,188 (18.6%) | 575 (21.1%) | 371 (22.1%) |
| | Quintile 2 | 418,408 (18.5%) | 30,277 (19.9%) | 531 (19.5%) | 358 (21.3%) |
| | Quintile 3 | 426,868 (18.8%) | 29,974 (19.7%) | 505 (18.6%) | 321 (19.1%) |
| | Quintile 4 | 437,204 (19.3%) | 26,305 (17.3%) | 502 (18.4%) | 286 (17.0%) |
| | Quintile 5 (most privileged) | 459,976 (20.3%) | 30,453 (20.0%) | 599 (22.0%) | 336 (20.0%) |
| | Unknown | 101,407 (4.5%) | 6,747 (4.4%) | 10 (0.4%) | 7 (0.5%) |

[a]Of 1,679 *Salmonella* isolates available for sequencing, 1,654 (99%) had an allele code assigned, 5 (<1%) underwent whole genome sequencing but failed quality control, and 20 (1%) were unique sequences that could not be matched to an existing allele code by CDC's naming algorithm.

[b]This index compares the prevalence of high income (≥$150,000) non-Hispanic white households vs. low income (<$30,000) people of color households, by census tract of patient residence, per Census 2010 boundaries, according to U.S. Census American Community Survey, 2015–2019 (11).

**Web Table 2.** Analysis conducted on June 15, 2023 to apply prospective tree-temporal scan statistics to detect emerging SARS-CoV-2 variants in specimens collected among New York City residents during the 14–28-day period ending May 31, 2023.

| Variant | No. of cases with specimens collected during 12-week study period, March 9–May 31, 2023 | Cluster start date, ending May 31, 2023 | No. of cases with specimens collected during cluster window | No. of expected cases | Relative risk | No. of excess cases | Test statistic | Recurrence interval (years) | No. of consecutive weeks signaling | Percent of sequenced cases with specimens collected week ending May 31, 2023 |
|---|---|---|---|---|---|---|---|---|---|---|
| XBB.1.16[a] | 164 | May 4 | 100 | 39.3 | 5.9 | 82.9 | 33.5 | 19,231 | 10 | 20 |
| XBB.2.3[a] | 64 | May 18 | 21 | 5.3 | 6.0 | 17.5 | 13.4 | 19,231 | 2 | 15 |
| XBB.1.5.16[a] | 10 | May 16 | 7 | 1.1 | 18.8 | 6.6 | 6.9 | 11 | 1 | <1 |
| XBB.1.9.2[a] | 75 | May 8 | 32 | 15.4 | 3.0 | 21.4 | 6.8 | 10 | 3 | 5 |
| XBB.1.5.39 | 16 | May 4 | 12 | 3.8 | 9.7 | 10.8 | 5.5 | 1 | 1 | 10 |

[a]Subvariants included.

### Web Appendix References
